# Day-to-Day Circadian Phase Fluctuations Shape Sleep and Behavior in Adolescents with ADHD

**DOI:** 10.64898/2026.04.30.26352043

**Authors:** Natacha Reich, Andrea Imparato, Maude Schneider, Stephan Eliez, Christopher Graser, Corrado Sandini

## Abstract

Sleep–wake regulation arises from the interaction between homeostatic sleep pressure and circadian timing, yet current assessments evaluate these processes independently and fail to capture their dynamic modulation by environmental pressures. This limitation is particularly relevant in adolescents with attention-deficit/hyperactivity disorder (ADHD), who are at increased risk of circadian delay and sleep disruption.

Here, we combined month-long wearable-based physiological monitoring with ecological behavioral assessments in adolescents with ADHD to characterize circadian and homeostatic processes dynamically in real-world settings. Using continuous skin temperature recordings, we derived individualized and day-specific estimates of circadian phase through hierarchical modelling, and integrated these measures with actigraphy-based sleep estimates and daily assessments of neurocognitive functioning and functional impairment.

Temperature-derived circadian phase correlated with questionnaire-based chronotype but more accurately predicted sleep patterns. Delayed circadian phase was associated with later sleep onset and greater weekday–weekend variability. Importantly, circadian phase exhibited significant day-to-day fluctuations, particularly in individuals with delayed phase, reflecting interactions with environmental constraints. Sleep latency was jointly determined by homeostatic sleep pressure and day-specific circadian phase, with combined models outperforming either process alone.

Crucially, both sleep deprivation and day-specific circadian misalignment independently predicted fluctuations in ADHD symptom severity, perceived stress, and neurocognitive impulsivity. In contrast, mean circadian phase alone did not explain behavioral variability.

These findings demonstrate that circadian regulation is a dynamic, environmentally sensitive process rather than a fixed trait. Wearable-based estimation of circadian phase provides a scalable approach to capture these dynamics and may enable personalized interventions targeting sleep and circadian dysregulation.

## Introduction

Sleep–wake regulation emerges from the interaction of two partially independent biological processes: a homeostatic process (Process S), which reflects the accumulation and dissipation of sleep pressure[1], and a circadian process (Process C)[2, 3], which regulates the daily timing of sleep and wakefulness [4]. Sleep onset typically occurs when sleep pressure is sufficiently high and circadian signals favor sleep.

The circadian system exerts its influence through rhythmic modulation of multiple physiological processes, including hormone secretion, alertness, and thermoregulation[2]. Body temperature is one of the most robust circadian outputs: core temperature decreases in the evening and night in close temporal association with melatonin secretion[5], while skin temperature shows an inverse pattern[6], increasing at night as melatonin induced peripheral vasodilation[7, 8], facilitates heat loss and promotes sleep initiation[9]. Because of this tight physiological coupling, skin temperature provides a non-invasive marker of circadian timing that can be measured continuously in naturalistic settings[6].

In clinical practice, however, circadian timing is most often inferred indirectly through chronotype questionnaires, which offer only a coarse and static approximation of an individual’s circadian phase[10-12]. In the academic setting, current gold standard estimation of circadian processes consists in measuring timing of melatonin production under strictly controlled laboratory conditions typically over 1 or 2 nights of recording[13]. Both approaches, however, overlook two central features of circadian regulation.

First, while current assessment measures homeostatic and circadian processes independently of each other, the two processes likely interact to perpetuate circadian dysregulation[14, 15] and modulate its downstream behavioral consequences[16]. For instance, increasing sleep pressure through daytime sleep restriction is a key component of behavioral interventions targeting circadian dysregulation[17-19], the effectiveness of which is comparable to that of pure modulation of circadian processes through melatonin supplementation[20, 21]. Moreover, sleep pressure alterations resulting from sleep disruptions could account for a significant proportion of behavioral consequences of circadian dysregulation (i.e. excessive morning sleepiness)[22, 23].

Second, current assessment approaches typically assume circadian rhythms to be fixed for each individual and thus conceptualize environmental factors that might modulate circadian redouts as confounds that need to be controlled for[10-13]. While the high-heritability of circadian rhythms would support the validity of this model[24, 25], the rapid epidemiological reduction in sleep duration[26-30], mirrored by societal changes in environmental modulators of circadian processes[28, 31, 32], would argue against a purely genetic pathophysiology of circadian dysregulation[2]. Of note, the effects of genetic vulnerability to circadian dysregulation might actually increase under more challenging environmental conditions[33-37], suggesting that circadian disturbances might be best understood as emerging from differential vulnerability to dynamically fluctuating exogenous modulators[2], rather than fixed endogenous processes that can be measured in a highly controlled laboratory vacuum.

Collectively, these results support the need to move beyond static and retrospective assessments of static circadian rhythms to a dynamic assessment of how circadian and homeostatic processes fluctuate and interact as functions of evolving contextual and environmental factors, which could represent a key stepping stone towards improving the effectiveness of interventions targeting sleep and circadian disturbances.

The clinical need to assessment and management of circadian dysregulation is particularly urgent for adolescents suffering from mental health and neurodevelopmental disorders such as ADHD[38, 39]. Indeed, adolescents undergo a striking change in circadian rhythm quantified as a 2.5 hour shift on average[40-42], and corresponding to approximately 1.5 standard deviations of a typical between-subject circadian-rhythm variability observed in adults[42]. Put differently, an adolescent with median circadian rhythm would correspond to a circadian rhythm observed at the 93^th^ percentile of an adult distribution or the effect of traveling across 3 time-zones (from Lisbon to Istanbul or from Los Angeles to Boston). Given that society largely fails to adapt to this biological shift[43], it is estimated that at least 50% of adolescents meet criteria for clinically significant sleep dysregulation[44].

The presence of ADHD exacerbates this worrisome picture[45]. Indeed, converging evidence has demonstrated a genetic and clinical association[46] between ADHD and circadian dysregulation with an overrepresentation of delayed circadian rhythm[47] that could contribute to explaining the overrepresentation of sleep disturbances (such as difficulties in initiating sleep or excessive daytime somnolence) observed in ADHD[48, 49]. Interestingly a converging body of evidence demonstrates that sleep dysregulation, resulting in excessive sleep pressure can directly modulate ADHD symptomatology[50-56] including both inattention[57] and impulse control deficits[58], that can be transiently induced by manipulating sleep in neurotypical individuals [59]. These considerations have raised the concern that sleep dysregulation resulting from exacerbation of circadian disruptions could contribute to a proportion of “delayed onset” cases of ADHD that are diagnosed or decompensate during adolescence[60]. While this question carries profound clinical and societal relevance it remains largely unanswered[61], due to limitations of current assessment approaches detailed above[62, 63]. Indeed, static retrospective measures do not allow to explicitly quantify the extent to which impairment in functioning observed in adolescents with ADHD can be attributable to circadian dysregulation and resulting sleep disturbances[64].

In the present study, we addressed this gap by combining month-long smartwatch-based physiological monitoring with smartphone-based self-reports in adolescents with ADHD. Using continuous skin temperature recordings, we derived individualized estimates of circadian phase while simultaneously capturing day-to-day fluctuations in circadian timing. By integrating these physiological measures with actigraphy-based sleep estimates and ecological momentary assessments of daily functioning, we examined how circadian phase relates to sleep–wake patterns and how circadian misalignment under weekday constraints shapes symptom expression across the week.

## Results

### Circadian phase estimation and validation

Multimodal behavioral phenotyping data across a variety of biological measures were collected using the Empatica smartwatch device for a total of 1192 days in 39 adolescents with ADHD (average number of nights per subject: 30.57 ± 9.95).

In the present analyses, we focused primarily on skin temperature, which is considered a proximal marker of circadian fluctuations. Data were averaged across 1-hour to improve the signal-to-noise ratio.

Skin temperature was then modelled as a 24-hour sine function, defined by phase, amplitude, and intercept, which explained a significant proportion of temperature signal variation (AIC = 4119.27; decrease of 1653.02 relative to a constant null model; LR = 1657.02, p<0.001, parametric bootstrap). This confirms a significant contribution of 24-hour circadian processes to temperature fluctuations. Including subject-specific random effects for phase, amplitude, and intercept significantly improved model fit (AIC = 38906.03; decrease of 2513.23 relative to the constant sinusoidal model; LR = 2525.23, p=0.0015, parametric bootstrap), consistent with substantial between-subject variability in underlying circadian processes. Notably, including day-specific random effects on top of subject-level terms contributed a further significant improvement in model fit (AIC = 37525.85; decrease of 1380.18 relative to the subject-level model; LR = 1392.18, p<0.001, parametric bootstrap), suggesting meaningful day-to-day variation in circadian processes per subject.

We next explored the external validity of this model, starting with validation of subject-specific effects. Subject-specific temperature phase (TP), capturing the extent to which an individual’s phase was advanced or delayed relative to the population average (see Figure 1.1.A), correlated significantly with the gold-standard retrospective estimate derived from the Horne–Östberg Morningness–Eveningness Questionnaire (MEQ) (Spearman’s ρ = 0.44, p = 0.03, see Figure 1.1.B), indicating that individuals with later temperature phase reported more evening-oriented chronotype preferences.

**Figure 1:**
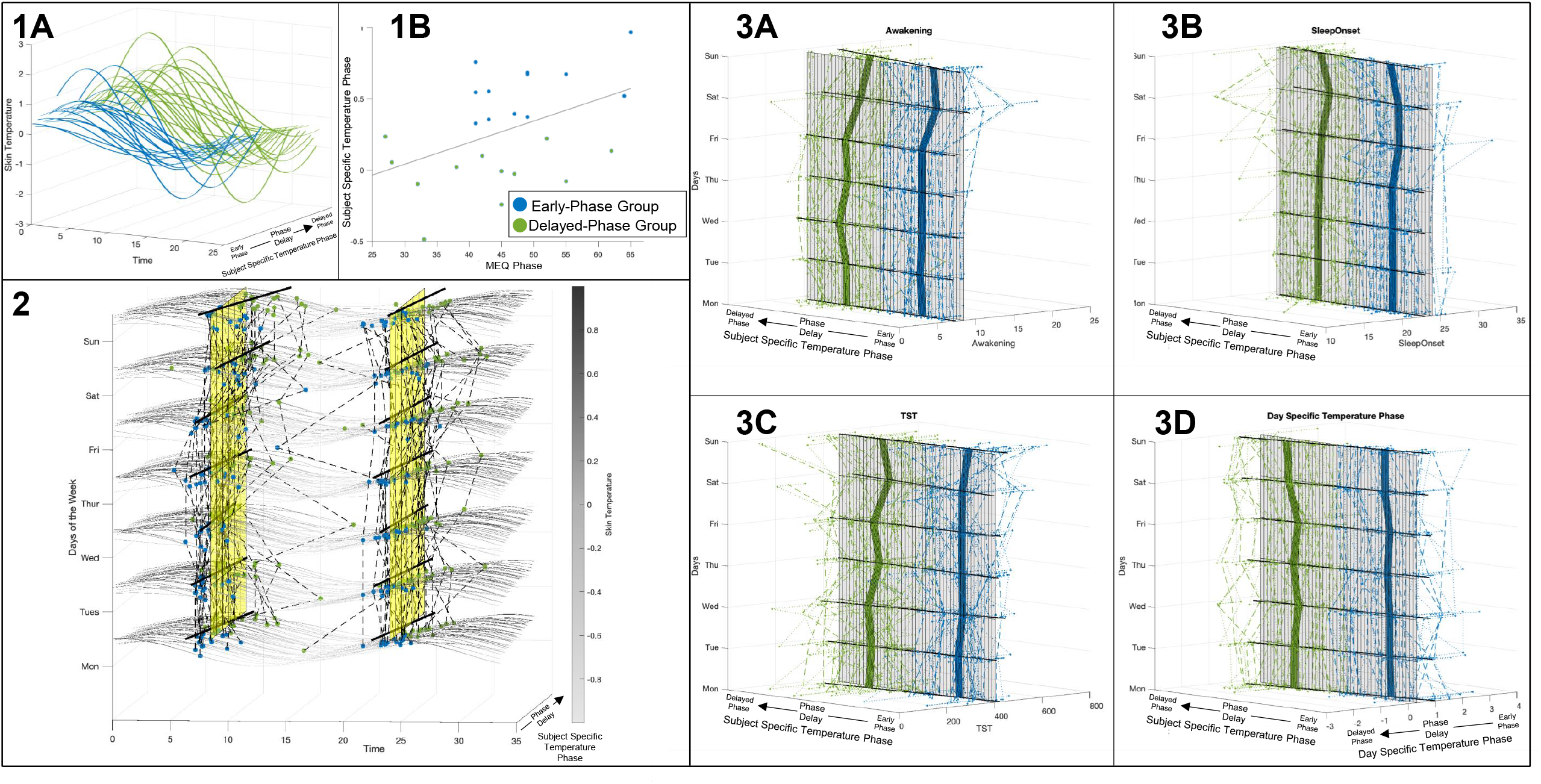
Dynamic circadian phase, temperature rhythms, and sleep timing across individuals and weekdays. **Panel 1: Validation of subject-specific circadian phase A**. Modelled 24-hour skin temperature rhythms. The x-axis represents clock time (hours), the y-axis shows participants ordered by their subject-specific temperature phase, and the color (z-axis) represents modelled wrist temperature. **B**. Association between subject-specific temperature phase (y-axis) and chronotype as assessed by the MEQ questionnaire (x-axis). **Panel 2**: **Day-specific temperature dynamics across the week**. Modelled temperature profiles are shown across a 35-hour window (x-axis: time of day), for participants ordered by their mean circadian phase (y-axis), and across days of the week (z-axis). Grey lines represent modelled skin temperature, for each participant and each day of the week. The yellow surfaces indicate the average sleep onset and awakening times across the full sample. Dots represent mean sleep onset and awakening times for each weekday. Black lines represent the association between subject-specific phase and sleep timing (sleep onset or awakening) for each day of the week. **Panel 3: Sleep variables and day-specific phase across weekdays**. For each participant (ordered by subject-specific phase; x-axis), sleep variables and day-specific temperature phase are plotted across days of the week (z-axis). The y-axis represents the variable of interest. Surfaces represent the association between circadian phase and each variable, while lines show day-specific regression fits. Individual trajectories are displayed for each participant, and group-level trends are overlaid. **A**. Awakening time **B**. Sleep onset **C**. Total sleep time **D**. Day-specific temperature phase (DSTP)

### Circadian modulation of sleep patterns

Having established external validity, we evaluated whether subject-specific temperature phase provided additional value in predicting downstream behavioral correlates of circadian variation, particularly sleep patterns, compared to the MEQ.

Sleep patterns were estimated from motion sensors using the Empatica algorithm, yielding dichotomous sleep/wake estimates at 1-minute resolution. From these data, we derived sleep onset timing, awakening time, and total sleep time (TST) across 745 nights. Subject-specific temperature phase (SSTP) was significantly correlated with average sleep onset timing (ρ=-0.34, p=0.03, N=39) and awakening time (ρ=-0.41, p=0.01, N=39). MEQ scores were not significantly correlated with either sleep onset timing (ρ=-0.35, p=0.09, N = 24) or awakening time (ρ=-0.08, p=0.7, N = 24). Supplementary analyses indicated that SSTP significantly predicted sleep onset (LR = 7.44, p = 0.006) and provided a better fit than MEQ (LR = 5.70, p = 0.017). Importantly, adding MEQ to the model including SSTP did not improve model fit (LR = 2.33, p = 0.13).

We next investigated whether average circadian phase moderated the impact of day-of-the-week–specific environmental factors on sleep patterns (see Figure 1.2). To this end, day of the week was modelled as a categorical factor in a linear mixed model.

Day-of-week factors significantly impacted all sleep variables, with the strongest effect observed for awakening time (LR = 161.70, p < .0001), which was delayed during weekends compared to weekdays. This delay contributed to longer TST during weekends (LR = 29.60, p < .0001), despite slightly delayed sleep onset (LR = 30.84, p < .0001).

Additionally, the effect of day of the week on awakening time was strongly moderated by SSTP, as indicated by a significant improvement in model fit when including SSTP × day-of-week interaction terms (awakening: LR = 22.19, p = 0.002). This interaction captured a greater tendency for delayed awakening during weekends in individuals with more delayed SSTP (Sun: t = -3.10, p-value = 0.004, see Figure 1.2 and 1.3.A). Hierarchical regression revealed that SSTP x day-of-week effects (vs null, LR = 134.15, p<0.0001) was superior to MEQ x day-of-week (vs null,, LR = 126.11, p<0.0001), in predicting delayed awakening and that MEQ x day-of-week did not significantly improve SSTP x day-of-week prediction accuracy (LR = 1.78, p = 0.97). These results suggest that SSTP is not only correlated with MEQ (supporting its external validity) but also outperforms MEQ in predicting downstream consequences of circadian phase variation across individuals.

As a post hoc analysis, we divided subjects into early and delayed circadian groups based on SSTP (Nearly = 18, Ndelayed = 21, see Figure 1.1.A) using k-means clustering. In the early-phase group, day-of-week fluctuations were limited to sleep onset (LR = 25.59, p = 0.0003) and awakening time (LR = 85.95, p < 0.0001) but did not impact TST (LR = 8.98, p = 0.17). The delayed-phase group showed significant fluctuations in sleep onset (LR = 15.20, p = 0.019), along with more pronounced fluctuations in awakening time (LR = 95.87, p < 0.0001), which contributed to significant variability in TST (LR = 30.18, p = 0.001).

Differences between groups were observed across most days for sleep onset, and primarily during weekends for awakening time (see supplementary table 1, see Figure 1.3.A and 1.3.B). The delayed circadian group also exhibited a fluctuating pattern of weekday TST reductions compared to the early-phase group (see Figure 1.3.C), which were particularly pronounced from Sun to Monday (t = -1,79, p=0.076), Tuesday to Wednesday (t = -2.62, p=0.01), and Thursday to Friday (t = -2.49, p=0.01). In contrast, Friday to Sat was the only interval during which TST showed a moderate increase in the delayed group relative to the early-phase group (t = 0.58, p=0.56).

### Dynamic circadian–homeostatic interactions

Having validated subject-level estimates of circadian phase, we next explored correlates of day-specific circadian phase estimates derived from our hierarchical modelling approach.

We first investigated whether day-specific TP estimates exhibited meaningful day-to-day fluctuations as a function of weekday-related environmental pressures. We found that day-specific-temperature-phase (DSTP) significantly fluctuated across weekdays (LR = 25.60, p<0.0001), and this effect was significantly moderated by SSTP (LR = 91.0, p < 0.001, see Figure 1.3.D).

Post hoc analyses revealed that, in the early-phase group, DSTP remained relatively stable across weekdays (LR = 7.60, p = 0.33). In contrast, the delayed-phase group exhibited significant fluctuations in DSTP (LR = 21.92, p = 0.002), characterized by increasing phase delay during the weekend, particularly on Sunday, which carried over into the early part of the week, followed by progressive phase advancement from Monday to Thursday (see Supplementary Table 1 and see Figure 1.3.D).

Next, we investigated whether DSTP fluctuations contributed to explaining day-to-day variability in sleep patterns, particularly sleep latency. Sleep latency (SL) was defined as the time to the next sleep epoch within a 120-minute window (values capped at 120 minutes), evaluated across successive 1-hour intervals from morning awakening until evening sleep onset.

We hypothesized that SL would be influenced by two independent mechanisms: (i) a homeostatic process reflecting progressive accumulation of sleep pressure during wakefulness, and (ii) a circadian process governing fluctuations in arousal, potentially captured by 24-hour temperature rhythms. Hour-specific sleep pressure estimates were modelled starting from awakening as the negative value of TST from the previous night and increased progressively during wakefulness, accounting for nap duration. Hour-specific temperature estimates were derived from modelled temperature using day-specific phase and amplitude parameters. SL was modelled as a function of time of day (1-hour bins from awakening to sleep onset), evaluating the added value of including sleep pressure and temperature, alone and in combination.

Sleep latency was significantly influenced by time of day, with a marked decrease during evening hours (LR 2514.77, p<0.0001), although substantial variability was observed both within and between subjects, particularly in the evening. Including sleep pressure by time-of-day interaction significantly improved model fit (LR = 207.29, p<0.0001, see Figure 2.1.A). Hour-specific temperature by time-of-day interaction also significantly improved SL prediction (LR = 443.73, P<0.0001, see Figure 2.1.B). Notably, modelling the combined effects of sleep pressure and temperature further improved model fit compared to modelling circadian (LR = 39.41, p=0.002) or homeostatic processes (LR = 275.85, p<0.0001) alone. In the evening (e.g., 22:00), both higher temperature (t = -4.34, p<0.0001) and higher sleep pressure (t = -1.98, p=0.049) independently contributed to shorter SL (see figure 2.2).

**Figure 2.**
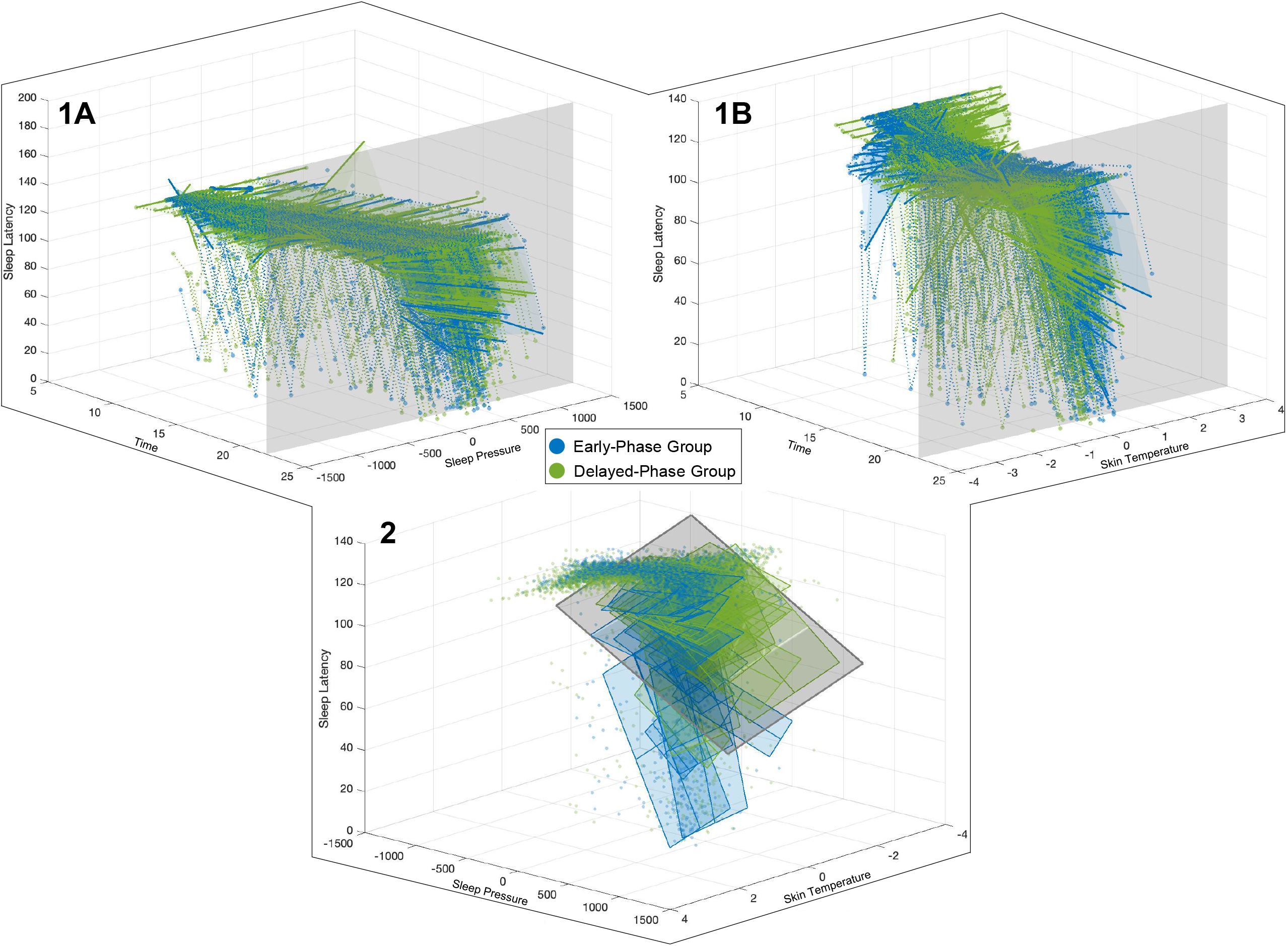
Dynamic interactions between circadian temperature, sleep pressure, and sleep latency. **Panels 1: Hourly associations between sleep latency, sleep pressure, and modeled skin temperature** These panels illustrate how sleep latency varies as a function of either sleep pressure (Panel A) or modeled skin temperature (Panel B) across the day. X-axis: Time of day (hours). Y-axis: Sleep pressure (Panel A) or modeled skin temperature (Panel B). Z-axis: Sleep latency. For each participant, a semi-transparent surface represents the modeled relationship between the predictor (sleep pressure or temperature) and sleep latency across the day, based on subject-specific mixed-effects models. Scatter points represent raw observations of sleep latency with corresponding sleep pressure or temperature values. Colored elements indicate circadian phase group (early vs delayed). Within each hour, regression lines are shown for each participant, capturing within-subject associations. A grey vertical surface at 22:00 highlights the specific time point examined in Panel C. **Panel 2: Joint effect of sleep pressure and circadian temperature on sleep latency (22:00)**. This panel shows the combined influence of sleep pressure and modeled skin temperature on sleep latency at 22:00. X-axis: Sleep pressure. Y-axis: Modeled skin temperature. Z-axis: Sleep latency. The grey surface represents the population-level model including both predictors. Colored semi-transparent surfaces represent individual participants, with color indicating circadian phase group. Scatter points correspond to raw data (sleep latency, sleep pressure, and temperature). Lines embedded within the surface indicate mean values of sleep pressure and temperature.

**Figure 3:**
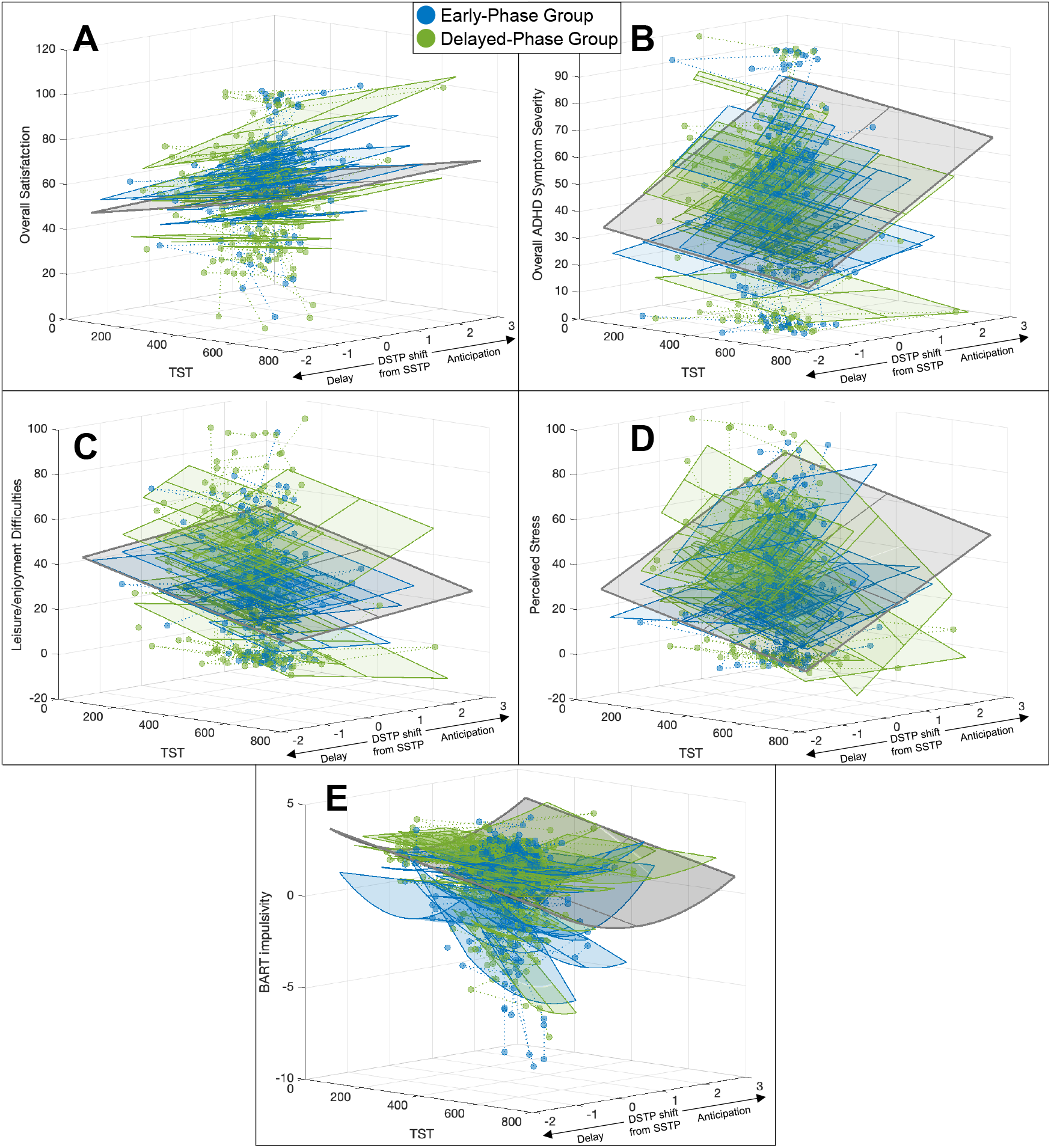
Joint effects of sleep duration and circadian misalignment on behavior and symptoms. This panel This figure illustrates how behavioral outcomes vary as a function of both Total Sleep Time (TST) and daily circadian phase deviation (i.e., the difference between daily phase [DSTP] and individual mean phase [SSTP]).X-axis: Total Sleep Time (TST) Y-axis: Daily phase deviation (DSTP – SSTP; positive values indicate phase anticipation) Z-axis: Behavioral outcome measure The grey surface represents the population-level mixed-effects model including both predictors (TST and phase deviation). Colored semi-transparent surfaces represent subject-specific model fits, with color indicating circadian phase group (early vs delayed). Scatter points correspond to raw observations (behavioral measures, TST, and phase deviation). Lines embedded within the surfaces indicate mean values of TST and phase deviation. Dashed trajectories represent within-subject changes across days. **Panel A**: Overall satisfaction **Panel B**: Overall ADHD symptom severity **Panel** C: Leisure/enjoyment difficulties **Panel D**: Perceived stress **Panel E**: BART impulsivity

Supplementary analyses revealed that a dynamic rolling-window approach to definition of day boundaries significantly improved model fit of sleep latency (at 22:00, T: t = -4.98, p<0.0001, SP: t = -3.34, p=0.0009, see supplementary material for description of the model). While initially introduced to control for potential reverse causal effects of sleep onset on temperature estimates, this approach also captured deviations from a perfect sinusoidal shape, revealing steeper temperature increases in the evening compared to morning declines. The fact that modelling such asymmetries improved SL prediction suggests that the steepness of evening temperature rise may reflect rapid increases in melatonin production, influencing both body temperature and arousal regulation.

To further disentangle the respective contributions of stable versus dynamic circadian influences on sleep initiation, we directly compared models including subject-level phase together with modelled temperature and sleep pressure at 22:00. Both sleep pressure and modelled temperature remained significant predictors after inclusion of SSTP (sleep pressure: t = −3.33, p = 0.001; temperature: t = −4.73, p < 0.0001), whereas SSTP itself was not associated with sleep latency (t = −0.93, p = 0.35) and did not improve model fit (LR = 0.8, p = 0.37). Together, these findings suggest that circadian influences on sleep initiation are more accurately characterized as dynamically fluctuating processes within individuals, rather than as fixed traits

We also tested whether day-to-day fluctuations in sleep pressure and circadian phase mediated the effects of day-of-week–specific environmental pressures on SL. Day-of-week significantly affected SL (LR = 24.37, p=0.0004, Fri: t=2.67, p=0.0078, Sat: t=2.21, p=0.028) sleep pressure (LR = 53.02, p<0.0001, Thurs: t=-2.51, p=0.012, Sat: t=-3.13, p=0.0018, Sun: t=-5.62, p<0.0001) and temperature (LR = 36.99, p<0.0001,, Fri: t=-3.5, p=0.0005, Sat: t=-4.23, p<0.0001) Importantly, day-of-week effects on SL were no longer significant after accounting for day-specific temperature and sleep pressure (Tues: t=-0.56, p=0.58, Wed: t=-1.66, p=0.1, Thurs: t=-0.63, p=0.53, Friday: t=1.84, p=0.07, Sat: t=0.80, p=0.42, Sun: t=-1.15, p=0.25), suggesting that environmental influences on sleep initiation are largely mediated by their influence on circadian and homeostatic processes.

### Behavioral impact of sleep and circadian variability

Finally, we investigated whether these fluctuations in sleep patterns impacted day-to-day variability in both neurocognitive difficulties and functional impairment in adolescents with ADHD.

Fluctuations in functional impairment across multiple domains were assessed using daily questionnaires (910 total questionnaires; 23.33 ± 12.14 per subject). These measures included overall ADHD symptom severity (“ADHD symptoms were a significant problem for me”), overall satisfaction (“I have the impression that today was a good day”), perceived stress (“I felt overwhelmed today”), and five domains of functional difficulty (academic, leisure/enjoyment, family relationships, peer relationships, and teacher relationships).

We also assessed neurocognitive fluctuations in cognitive control using a digital smartphone-based implementation of the Balloon Analogue Risk Task (D-BART) [65], administered a total of 1257 times (33.08 ± 25.69 per subject). We recently validated D-BART against the gold-standard laboratory-based CPT-3 neurocognitive battery, identifying a multivariate D-BART-Impulsivity response that differentiates adolescents with ADHD from typically developing controls and captures vulnerability to real-life behavioral consequences of impulse control deficits, with superior accuracy compared to CPT-3 assessments [66]. In the ADHD sample, we further observed significant within-subject fluctuations in BART-Impulsivity readouts. Here, we build upon these findings by investigating the impact of sleep patterns on day-to-day fluctuations in BART-Impulsivity scores.

We used forward stepwise regression models to predict fluctuations in both functional impairment domains and BART-Impulsivity scores as a function of multiple sleep variables, including TST (previous night), 2-night average TST, SSTP, DSTP, the difference between SSTP and DSTP, the square of the difference between SSTP and DSTP, and day of the week. Results of forward stepwise regression are reported in the supplementary table 2.

Regarding functional impairment domains, we found that day of the week was the only significant predictor of academic difficulties (Δ_BIC_: 24.6, LR = 59.92, p<0.0001, Sat: t = -5.99, p<0.0001, Sun: t = -4.89, p<0.0001) and teacher-relationship difficulties (Δ_BIC_: 19.69, LR = 55.01, p<0.0001, Sat: t = -5.09, p<0.0001, Sun: t = -4.98, p<0.0001), which remitted during weekends, Reduced TST predicted leisure/enjoyment difficulties (Δ_BIC_: 6.69, LR = 12.58, p=0.0004, t = -3.58, p=0.0004, see figure 2.C) and perceived stress (Δ_BIC_: 8.7, LR = 14.59, p=0.0001, t = -3.38, p=0.0008, see figure 2.D), while 2-night average TST predicted overall satisfaction (Δ_BIC_: 4.33, LR = 10.22, p=0.001, t = 3.22, p=0.001). Anticipation of DSTP relative to SSTP predicted higher overall ADHD symptom severity (Δ_BIC_: 3.28, LR = 9.17, p=0.002, t = 3.05, p=0.0025, see figure 2.B) and higher perceived stress (Δ_BIC_: 0.18, LR = 6.07, p=0.01, t = 2.47, p=0.01, see figure 2.D), even after accounting for TST.

Increased BART-Impulsivity was predicted by a combination of reduced TST (Δ_BIC_: 2.86, LR = 9.47, p=0.002, t = -3.09, p=0.002, see figure 2.E) and the DSTP–SSTP phase difference (t = -4.01, p<0.0001) together with the square of the phase difference (Δ_BIC_: 7.65, LR = 20.86, p<0.0001, t = 4.15, p<0.0001, see figure 2.E), suggesting that impulsivity increased both when DSTP was advanced and when it was delayed relative to SSTP, and tended to normalize when DSTP aligned with SSTP.

To our knowledge, this study is the first to comprehensively characterize day-to-day fluctuations in both homeostatic and circadian processes and to disentangle their contributions to functional impairment in adolescents with ADHD. Our results suggest that fluctuations in both homeostatic and circadian sleep processes directly predict the severity of neurocognitive and behavioral difficulties, accounting for a significant proportion of both between-subject variance and within-subject day-to-day variability.

## Discussion

In this study, we aimed to characterize circadian and homeostatic processes in a dynamic and ecologically valid manner, and to examine their contribution to sleep patterns and behavioral functioning in adolescents with ADHD.

Firstly, we show that we can estimate subject-specific circadian phase using advanced modelling of skin temperature signals obtained from a highly scalable wearable device. The resulting readout was correlated with gold-standard questionnaire measures but provided superior accuracy in predicting the biologically relevant impact of average circadian phase on sleep patterns. In particular, we show that this approach captures across-subject variability in sleep patterns, including the impact of delayed circadian phase on sleep initiation difficulties, as well as the tendency for more erratic weekday fluctuations in sleep patterns, stemming from the dissociation between endogenous circadian phase and exogenous environmental pressures. This, in and of itself, could prove clinically useful by providing an objective readout of social jetlag that outperforms current circadian rhythm estimation approaches based on either retrospective evaluation or objective analysis of sleep patterns[6, 11, 12, 67, 68]. Indeed, both the subjective experience of arousal fluctuations and the objective analysis of sleep pattern fluctuations are not only impacted by circadian rhythm but are also tightly shaped by independent homeostatic sleep processes[69]. Our proposed temperature-based estimation represents a significant advancement in this regard, as it allows estimation of circadian rhythm based on a biological skin temperature signal that is both a direct and intrinsic correlate of circadian fluctuations[8, 9] and relatively independent of homeostatic sleep processes[4].

A second key innovation provided by our approach is the ability to move from a static characterization of average circadian rhythm to a dynamic view of within-subject, day-to-day variation in circadian processes. While current assessment approaches typically conceptualize circadian phase as a fixed subject-specific trait[10-13], converging lines of evidence demonstrate that circadian processes are tightly modulated by a variety of dynamically fluctuating behavioral and environmental factors[2], most notably light exposure[32]. The ability to capture the impact of such exogenous modulators carries substantial clinical relevance, given overwhelming epidemiological evidence that societal factors play a prominent role in shaping sleep patterns[28, 31, 32], and that adolescents may be particularly vulnerable to such pressures[44]. In line with the influence of these exogenous modulators, our findings directly demonstrate that circadian phase does not only act as a fixed trait moderating the effects of environmental pressures but is also dynamically shaped by them. Subject-specific factors remain relevant but act primarily as moderators of exogenous influences on day-specific circadian phase, which appear to be particularly important in individuals with more delayed average circadian phase. The consequences of circadian delay were not limited to the social jetlag captured by recurrent endogenous–exogenous misalignment but also included more pronounced and recurrent day-to-day misalignments between an individual’s own endogenous circadian phases. This phenomenon can be likened to the effects of repeated travel across time zones[70] or shift work [71-73], which are not limited to recurrent social jetlag, but also require continuous realignment of endogenous circadian phase in response to changing environmental pressures.

A third major contribution of this study is the ability to model the interaction between circadian and homeostatic processes in naturalistic settings. While experimental paradigms typically isolate these processes, real-world sleep regulation emerges from their continuous and bidirectional interaction[14, 15, 69]. Our results confirm that both processes independently and jointly predict sleep initiation, with combined models providing superior explanatory power. Notably, the effects of day-of-week–specific environmental factors on sleep latency were fully accounted for by their impact on day-specific circadian phase and sleep pressure. Furthermore, day-specific circadian estimates outperformed subject-level averages in predicting sleep patterns, supporting the view that circadian regulation is best understood as a dynamic process rather than a fixed trait. While the role of sleep pressure in sleep initiation is well established[1, 74], the demonstration that day-to-day fluctuations in circadian phase independently shape sleep propensity represents a novel and clinically relevant insight. From a research perspective, these findings open new avenues for characterizing not only average circadian timing but also individual vulnerability to environmental modulation. Such vulnerability may provide a more sensitive phenotype for investigating the biological and genetic underpinnings of circadian dysregulation[34, 36, 37]. Moreover, the ability to track dynamic circadian fluctuations could pave the way for personalized intervention strategies that optimize behavioral and pharmacological treatments based on objective readouts of their impact on circadian processes. For instance, the effects of melatonin depend on endogenous circadian timing[75], and tailoring administration to individual phase dynamics could improve effectiveness[76]. Similar considerations apply to a wide range of interventions whose effects may be modulated by circadian phase[77], including chemotherapy[78].

A fourth and final contribution of our analytical approach was the ability to dissect the role of dynamic circadian and homeostatic sleep fluctuations in the day-to-day expression of ADHD neurocognitive and behavioral symptomatology. Converging evidence suggests that the adolescent predisposition to circadian delay and reduced sleep duration is particularly pronounced in individuals with ADHD[45, 47-49, 60]. In parallel, experimental studies have shown that sleep alterations of similar magnitude can induce behavioral changes resembling ADHD, including executive dysfunction[59, 79-81], impaired impulse control[82], and emotional dysregulation[83-86]. However, quantifying the contribution of sleep alterations to behavioral difficulties in ADHD remains challenging, as it requires dynamic assessment of both sleep and symptom fluctuations[61]. Here, we leverage recent advances in digital phenotyping to address this gap. We show that fluctuations in homeostatic sleep pressure, resulting from recurrent sleep deprivation, play a key role in predicting the severity of both neurocognitive impulse control deficits and key domains of functional impairment, particularly overall daily satisfaction, reduced enjoyment of leisure activities, and higher perceived stress. These results align with a large body of research highlighting the impact of sleep deprivation and increased sleep pressure on neurocognitive and behavioral functioning, especially inhibitory control[82] and emotional regulation [83-86]. Importantly, our dynamic multimodal phenotyping approach advances prior cross-sectional work by directly capturing the impact of sleep fluctuations on within-subject neurocognitive and behavioral variability. This ability to quantify subject-specific sensitivity to sleep alterations may have important clinical implications, supporting more individualized management strategies based on personal sleep needs rather than population averages[62, 63].

Finally, in addition to characterizing the role of homeostatic sleep pressure, our results provide novel evidence supporting the importance of moving from static to dynamic characterization of circadian rhythm fluctuations. Day-specific circadian phase estimation was not only relevant for predicting fluctuations in sleep patterns but also represented an independent predictor of fluctuations in overall ADHD symptom severity and associated levels of perceived stress. The impact of these circadian phase fluctuations was not limited to subjective evaluations of behavioral symptoms but also contributed to explaining objective day-to-day fluctuations in neurocognitive impulsivity, even when accounting for the significant effects of sleep deprivation on cognitive control deficits. In contrast, mean circadian phase alone did not provide additional predictive value, suggesting that circadian dysregulation may be better understood as a spectrum of dynamic fluctuations influenced by environmental pressures, resulting in varying degrees of misalignment between subject-level and day-specific circadian phase[2]. Such day-specific endogenous misalignment may represent a novel biological readout more proximally linked to downstream neurocognitive performance, mediating the effects of both endogenous circadian traits and exogenous environmental influences. These findings align with a substantial body of basic and clinical evidence indicating that manipulation of circadian phase has biological and behavioral effects that are not fully explained by associated sleep alterations[70-73]. Our approach provides a novel quantitative framework to investigate these circadian dynamics in real-world settings. This framework may have broader relevance for understanding symptom variability across a range of psychiatric conditions associated with circadian disruption, including bipolar disorder[87], conduct disorders[88], addiction disorders[89] and emotional dysregulation disorders[90]. Notably, impulse control deficits—directly associated with circadian misalignment in our findings—represent a common behavioral feature across these disorders[91-97], which also show increased prevalence during adolescence[89, 98, 99], coinciding with adolescent increase in impulsivity [100, 101] and circadian phase delay[40]. Finally, the ability to quantify these dynamic circadian fluctuations may have implications beyond psychiatry[77], given the growing recognition of circadian influences on metabolic[102], cardiovascular[103], and oncological processes[78].

Several limitations should be acknowledged. First, while skin temperature provides a validated proxy for circadian phase, it remains an indirect measure and may be influenced by external factors such as ambient temperature or physical activity. Although our modelling approach mitigates some of these effects, future studies incorporating direct measures of melatonin would strengthen validation. Second, the sample size and focus on adolescents with ADHD may limit generalizability. Replication in larger and more diverse populations will be important. Third, while observational design enhances ecological validity, it precludes causal inference, and experimental studies will be needed to confirm directionality.

In conclusion, our findings demonstrate that circadian and homeostatic sleep processes can be dynamically characterized in real-world settings and that their interaction plays a key role in shaping sleep patterns and behavioral functioning in adolescents with ADHD. These results support a shift from static to dynamic models of circadian regulation and highlight the potential for personalized, mechanism-based approaches to the assessment and treatment of sleep and circadian disturbances.

## Methods

### Participants

In the study, 39 participants with idiopathic ADHD (mean age = 15.52 ± 1.9 years, M/F= 22/19) were recruited shortly after receiving their diagnosis. All participants, and legal guardians, when applicable, provided written informed consent in accordance with institutional ethical guidelines.

### Study Protocol

Participants completed a 30-day ecological monitoring protocol designed to capture sleep, circadian rhythms, and daily functioning under naturalistic conditions. Passive physiological data were collected continuously using wearable devices, while active self-report data were collected via smartphone-based surveys.

### Wearable Sensing

Participants wore an Empatica EmbracePlus (https://www.empatica.com/embraceplus/) smartwatch throughout the monitoring period. The device is equipped with accelerometers and gyroscopes to assess movement, photoplethysmography sensors to estimate heart rate and heart rate variability, thermistors to measure skin temperature, and additional sensors to estimate respiratory rate and oxygen saturation. The present analyses focused on skin temperature and movement-derived sleep estimates. Sleep–wake patterns were inferred using validated actigraphy-based algorithms that estimate sleep probability from movement data (Regalia et al., 2021).

### Ecological Momentary Assessments

Participants completed ecological momentary assessments (EMA) delivered via smartphone six times per day, as well as a morning and evening survey. These assessments captured momentary mood, subjective sleep quality, ADHD symptoms, daily difficulties, and overall functioning. This intensive sampling allowed examination of daily and weekly symptom fluctuations in relation to circadian timing and sleep–wake behavior.

### Neurocognitive assessment

Neurocognitive impulsivity was assessed using a smartphone-based implementation of the Balloon Analogue Risk Task (D-BART)[65], administered repeatedly in daily life. This paradigm provides an ecologically valid measure of impulsive decision-making and cognitive control. A detailed description of the paradigm administration and analysis pipeline is detailed elsewhere [66] and described briefly here. Specifically, a multivariate latent dimension of impulsivity was identified using partial least squares (PLS) analysis linking D-BART performance to gold-standard laboratory-based neurocognitive measures (CPT-3). This latent component captured a shared pattern of variance reflecting individual differences in neurocognitive impulsivity responding across tasks.

In the present study, we quantified neurocognitive impulsivity by projecting trial-level D-BART performance onto this previously validated latent component. Specifically, D-BART variables were first standardized using the scaling parameters derived from the original PLS model and then weighted by the corresponding PLS loadings to compute a single composite score for each assessment. This score reflects the extent to which an individual’s behavior aligns with the neurocognitive impulsivity profile identified in the reference dataset, with higher scores reflecting increased neurocognitive impulsivity.

### Chronotype Questionnaire

Chronotype preference was assessed using the Morningness–Eveningness Questionnaire (MEQ; Horne & Ostberg, 1976). MEQ total score was used to validate temperature-derived estimates of circadian phase.

### Circadian Modeling of Skin Temperature

Circadian rhythmicity in skin temperature was characterized by fitting a sinusoidal model with a fixed 24-hour period to each participant’s temperature data. This approach captures the fundamental rhythmic structure of circadian physiology while allowing individual differences in timing and amplitude.

Temperature at time *t* was modeled as:

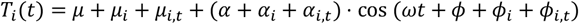

where μ represents the overall mean temperature, α the rhythm amplitude, and ϕ the circadian phase. Participant-level (*i*) and day-level (*i,t*) random effects captured stable inter-individual differences and day-to-day variability.

To enable linear mixed effects estimation, this model was transformed to the following form

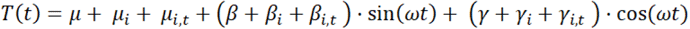

from which the circadian amplitude and phase terms (A, ^Ф^)form the initial equation can be recovered as 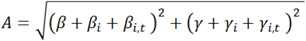 and Ф = *atan*2(*β* + *β*_*i*_ + *β*_*i,t*_, *γ* + *γ*_*i*_ + *γ*_*i,t*_) respectively.

#### From these models, we derived

Subject-specific-temperature-phase (SSTP): reflecting stable individual circadian timing Day-specific-temperature-phase (DSTP): capturing within-individual daily fluctuations in circadian phase

### Sleep Onset and Awakening Identification

Sleep–wake states were recorded in 1-minute epochs using wearable-derived sleep staging. Because the device did not provide explicit timestamps for sleep onset or final awakening, these events were identified algorithmically from transitions in the epoch-level sleep–wake data.

#### Sleep Onset

Sleep onset was defined as the first transition from sustained wakefulness to sustained sleep within a night. A candidate transition was retained as a valid sleep onset if:

- At least 50% of the preceding 60 minutes were classified as wake, and
- At least 50% of the subsequent 60 minutes were classified as sleep (including brief awakenings and sleep interruptions).

This stability criterion reduced the influence of short or unstable sleep attempts. When multiple sleep transitions occurred within a short period, only the earliest stable transition was retained. If a transition occurred adjacent to a short period of missing data, sleep onset was estimated as the midpoint between the last valid wake epoch and the first valid sleep epoch, provided the missing segment did not exceed 60 minutes.

#### Awakening

Final awakening was defined analogously as the transition from sustained sleep to sustained wakefulness. A transition was classified as a valid awakening if:

- At least 50% of the preceding 60 minutes were classified as sleep, and
- At least 50% of the subsequent 60 minutes were classified as wake.

Short awakenings followed by rapid return to sleep were excluded. When awakening occurred near missing data segments, the midpoint between the last sleep epoch and the first wake epoch was used if the missing period was shorter than 90 minutes.

#### Night Definition and Episode Selection

When multiple sleep episodes occurred within the same circadian cycle, the episode with the longest total sleep duration was selected as the main nocturnal sleep period.

### Total sleep time was then computed for each night

In all analyses, sleep onset was assigned to the calendar day preceding the corresponding awakening time (e.g., “Sun” sleep refers to the night from Saturday to Sunday).

#### Sleep pressure and Sleep Latency

Sleep latency was categorized as the time (in minutes) until the next sleep epoch within a 120-minute observation window, with values capped at 120 minutes when no sleep occurred.

Sleep pressure was estimated as the cumulative time spent awake since morning awakening, minus any sleep obtained during the day or night, such that sleep pressure increased with sustained wakefulness and decreased following sleep episodes.

Both measures were computed continuously from morning wake time to bedtime and were sampled at hourly intervals.

## Statistical Analyses

Skin temperature was modelled using a hierarchical 24-hour sinusoidal function characterized by phase, amplitude, and intercept parameters, in order to capture underlying circadian structure (see *Methods*: *Circadian Modeling of Skin Temperature)*. Relative model performance was evaluated by sequentially comparing nested models of increasing complexity: (i) constant sinusoidal model (vs null), (ii) a model including subject-level random effects on intercept and circadian parameters, and (iii) a model additionally including day-level random effects for each of these terms. For each comparison, likelihood ratio statistics were computed. Empirical p-values were obtained using parametric bootstraps, for which data were simulated according to the simpler model, both models were refitted to each simulated dataset, and significance was assessed by comparing the observed LR statistic to its bootstrap distribution. Bootstrap procedures were based on 1000 simulations per comparison; iterations for which the more complex model-estimation failed to converge were excluded. This occurred only for the comparison between models (ii) and (i), with a failure rate of approximately 32%. P-values were computed from successful replicates.

To assess the validity of circadian phase estimates, subject-specific temperature phase (SSTP) was correlated with self-reported chronotype measured using the Morningness–Eveningness Questionnaire (MEQ). Associations were evaluated using Spearman rank correlations. To further examine ecological validity, SSTP and MEQ scores were compared in their ability to predict sleep–wake behavior derived from actigraphy. Sleep onset time and awakening time were used as primary markers of sleep–wake patterns. Additionally, we compared the predictive value of SSTP and MEQ for sleep onset using likelihood ratio tests, by evaluating models including SSTP or MEQ separately (Sleep Onset ∼ SSTP; Sleep Onset ∼ MEQ), as well as a combined model including both predictors (Sleep Onset ∼ SSTP + MEQ).

To investigate whether average circadian phase moderates the impact of environmental constraints across the week, sleep outcomes (sleep onset time, awakening time, and TST) were modeled using linear mixed-effects models with day of the week specified as a categorical predictor (Monday as reference). Model comparisons were performed using likelihood ratio tests to evaluate the contribution of SSTP and its interaction with day of the week:

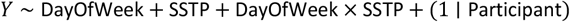

To further characterize day-specific effects, separate models were estimated for each day of the week, examining the association between SSTP and sleep outcomes:

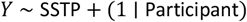

To compare the relative contributions of SSTP and MEQ in capturing day-of-week effects on sleep timing, we fitted mixed-effects models including interaction terms between day of the week and either SSTP or MEQ. Model comparisons were performed using likelihood ratio tests to assess improvements in fit relative to null models, as well as the added value of including MEQ alongside SSTP.

To identify subgroups with systematically different circadian timing, k-means clustering was applied to SSTP estimates, with the number of clusters fixed to two to distinguish relatively early and delayed circadian profiles (Nearly = 18, Ndelayed = 21).

Analyses were then repeated within each subgroup to examine the effect of day of the week on sleep outcomes:

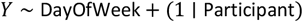

Finally, to further explore group differences at the daily level, day-specific models were estimated to assess the effect of circadian group on sleep outcomes for each day of the week:

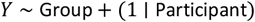

### Dynamic circadian–homeostatic interactions

To move beyond stable inter-individual differences and examine intra-individual variability, we investigated day-to-day fluctuations in circadian phase. Day-specific temperature phase (DSTP) was modeled using linear mixed-effects models to assess the influence of environmental constraints across the week. Specifically, we examined the effects of day of the week and subject-specific phase (SSTP), as well as their interaction, using likelihood ratio tests:

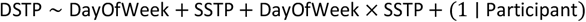

To characterize group-specific patterns, we then assessed the effect of day of the week on DSTP within circadian subgroups (early vs delayed):

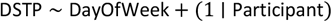

Finally, day-specific differences between circadian groups were examined using separate models for each day of the week:

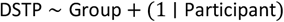

To investigate the joint influence of circadian (Process C) and homeostatic (Process S) mechanisms on sleep initiation, sleep latency (SL) was modeled as a function of skin temperature and sleep pressure across the day. Linear mixed-effects models included interactions with time of day (hourly bins), and model comparisons were conducted using likelihood ratio tests:

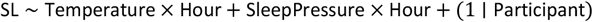

To further assess time-specific effects, additional models were estimated separately for each hour of the day:

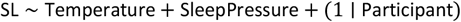

This approach allowed us to quantify the relative contributions of circadian and homeostatic processes at different times of day.

To account for potential temporal overlap between predictor and outcome, we repeated these time-specific models using temperature estimates derived from the dynamic rolling-window reconstruction described in the Supplementary Methods. This approach provides hour-specific temperature trajectories that minimize within-day circularity while preserving temporal alignment with sleep latency.

To disentangle stable versus dynamic circadian influences on sleep initiation, we focused on sleep latency at 22:00. Linear mixed-effects models were fitted including SSTP, modelled skin temperature, and sleep pressure as predictors of sleep latency (SL_(22:00)_ ∼ Temperature + SleepPressure + SSTP + (1 | Participant)). Model comparisons were conducted using likelihood ratio tests to evaluate the contribution of SSTP beyond temperature and sleep pressure.

Finally, we examined whether day-to-day fluctuations in circadian phase and sleep pressure mediated the effects of environmental constraints on sleep initiation, specifically at 22:00. We first tested the effect of day of the week on sleep latency, sleep pressure, and temperature using linear mixed-effects and model comparisons were conducted using likelihood ratio tests. We then assessed whether the inclusion of temperature and sleep pressure attenuated the effect of day of the week on sleep latency, indicating a potential mediating role of these physiological processes.

### Behavioral impact of sleep and circadian variability

We next examined whether day-to-day fluctuations in sleep and circadian processes predicted variability in neurocognitive functioning and functional impairment.

Daily functioning was assessed using repeated smartphone-based questionnaires administered across the study period (910 total questionnaires; 23.33 ± 12.14 per subject). Outcomes included overall ADHD symptom severity, perceived stress, overall satisfaction, and functional impairment across multiple domains (academic functioning, leisure/enjoyment, family relationships, peer relationships, and relationships with teachers).

Neurocognitive impulsivity was indexed using repeated D-BART–derived scores capturing a validated dimension of impulsive decision-making in daily life.

Daily fluctuations in questionnaire-based outcomes and neurocognitive impulsivity were modeled as a function of sleep and circadian predictors, including total sleep time (TST; previous night), 2-night average TST, subject-specific circadian phase (SSTP), day-specific circadian phase (DSTP), circadian misalignment (DSTP − SSTP) and its quadratic term (DSTP − SSTP)^2^, as well as day of the week.

Model selection was performed using a forward stepwise procedure based on the Bayesian Information Criterion (BIC) (see Supplementary Methods). At each step, candidate predictors were evaluated, and variables were retained only if they improved model fit relative to the previous model (i.e., reduction in BIC). All models included a random intercept for participant to account for within-subject dependence.

## Data Availability

All data produced in the present study are available upon reasonable request to the authors

## Acknowledgments

This study was supported by the Swiss National Science Foundation (SNSF; Grant No. 320030_212476 to SE and 10002308 to MS). Corrado Sandini was supported by an SNSF grant (No. 209096) and by the Fondation Gertrude von Meissner (Grant No. 52, March 2025). Natacha Reich received a scholarship from the Fondation Pôle Autisme. We warmly thank all the families who participated in the study.

