## Supplementary Material for "Day-to-Day Circadian Phase Fluctuations Shape Sleep and Behavior in Adolescents with ADHD"

### Methods

Time-resolved reconstruction of circadian temperature for sleep latency modelling

In our circadian modelling framework, participant-by-day random effects allow predicted temperatures at a given timepoint to depend strongly on observations from the same day. To avoid potential circularity in the sleep latency analyses, where temperature is used to predict sleep onset, we therefore reconstructed time-resolved temperature estimates using a modified approach that preserves temporal specificity while preventing overlap between predictor and outcome time windows.

Specifically, we used a two-step hour-shifted reconstruction procedure. First, we fit the hierarchical circadian model multiple times using 24 different definitions of the “day,” each time shifting the 24-hour boundary from 00:00 to 23:00. This yields a set of models in which circadian parameters are estimated under slightly different temporal segmentations of the data. Second, for each observation at clock time h, we extracted predicted temperature values from the model whose day boundary was defined immediately after that hour (i.e., using a shift of h + 1 hours). This procedure ensures that temperature estimates used to predict sleep latency are derived from model fits that do not incorporate information from subsequent observations within the same day, thereby minimizing temporal overlap between predictor and outcome.

Together, this approach yields hour-specific predicted temperature trajectories that respect the hierarchical structure of the model while providing temporally aligned estimates for sleep latency analyses.

#### Stepwise hierarchical model selection procedure

To identify sleep and circadian predictors associated with day-to-day fluctuations in behavioral and neurocognitive outcomes, we implemented a forward stepwise model selection procedure within a linear mixed-effects (LME) framework.

**Model specification**

For each dependent variable, models were specified with a random intercept for participant to account for within-subject dependence:

$$Y\sim\text{fixed effects}+(1\mid\text{Participant})$$

Candidate fixed effects included total sleep time, two-night averaged TST, day-specific circadian phase, subject-specific circadian phase, and the difference between day-specific circadian phase and subject-specific circadian phase, as well the square of the difference between SSTP and DSTP.

**Stepwise selection procedure**

Model selection proceeded iteratively, starting from a null model including only the random intercept:

$$Y\sim1+(1\mid\text{Participant})$$

At each iteration, all remaining candidate predictors not yet included in the model were tested independently by adding them to the current model. For each candidate variable, a new model was fit and compared to the previous model using the Bayesian Information Criterion (BIC) and likelihood ratio tests.

The predictor yielding the greatest improvement in model fit, defined as the largest reduction in BIC relative to the current model, was selected for inclusion, provided that:

1. The BIC of the updated model was lower than that of the previous model, and
2. The corresponding likelihood ratio test indicated a nominal improvement in fit (p < 0.05).

If no candidate predictor satisfied both criteria, the selection procedure was terminated.

After each inclusion step, the selected predictor was retained in the model, removed from the pool of candidate variables, and the procedure was repeated with the updated model.

**Model outputs**

For each dependent variable, the procedure yielded a final selected model, the sequence of retained predictors and for each inclusion step, the associated likelihood ratio statistic, p-value, and change in BIC (ΔBIC).

### Results

**Group differences in sleep and circadian measures across weekdays**

This supplementary table reports group-level differences between the early and delayed circadian phase groups across each day of the week. Mixed-effects linear regression models were used to compare groups on sleep onset time, awakening time, total sleep time (TST), and day-specific temperature phase (DSTP), with participant included as a random intercept.

|  | *Sun/Mon* | | *Mon/Tues* | | *Tues/Wed* | | *Wed/Thurs* | | *Thurs/Fri* | | *Fri/Sat* | | *Sat/Sun* | |
| --- | --- | --- | --- | --- | --- | --- | --- | --- | --- | --- | --- | --- | --- | --- |
|  | tStat | p-value | tStat | p-value | tStat | p-value | tStat | p-value | tStat | p-value | tStat | p-value | tStat | p-value |
| *Sleep Onset* | **3.24** | **0.002** | 1.85 | 0.07 | **2.12** | **0.037** | **2.54** | **0.01** | **3.35** | **0.001** | 0.76 | 0.45 | **2.02** | **0.047** |
| *Awakening* | 1.08 | 0.28 | **2.06** | **0.04** | -0.62 | 0.53 | 1.76 | 0.08 | 1.66 | 0.1 | 0.63 | 0.53 | **3.71** | **0.0003** |
| *TST* | -1.79 | 0.076 | -1.79 | 0.076 | **-2.62** | **0.01** | -0.45 | 0.65 | **-2.49** | **0.01** | 0.58 | 0.56 | -1.33 | 0.19 |
| *DSTP* | **-5.06** | **0.0001** | **-6.3** | **0.0001** | **-2.75** | **0.007** | **-3.85** | **0.0002** | **-4.39** | **0.0001** | **-4.85** | **0.0001** | **-4.92** | **0.0001** |

Supplementary Table 1 : Differences in sleep and circadian timing across the week.

*Differences in sleep variables between groups tested with Mixed Model Linear Regression across days of the week. For days, we report the specific tStat and p-values of group effect derived from the model. Significant effect are highlighted in bold.*

**Stepwise mixed-effects models predicting behavioral and neurocognitive outcomes**

We report the results of forward stepwise linear mixed-effects models used to predict day-to-day fluctuations in functional impairment domains and BART-Impulsivity scores. Candidate predictors included total sleep time (TST; previous night), 2-night average TST, subject-specific temperature phase (SSTP), day-specific temperature phase (DSTP), the difference between DSTP and SSTP, the square of the difference between SSTP and DSTP, and day of the week.

At each step, predictors were added based on improvement in model fit, assessed using likelihood ratio tests and ΔBIC. For each outcome, only predictors that improved model fit relative to the previous model were retained.

| **Outcomes** | **Predictors** | **LR** | **p-value** | **Δ_BIC_** |
| --- | --- | --- | --- | --- |
| Academic difficulties | Day of the week | 59.91 | 0.0001 | 24.6 |
| teacher-relationship difficulties | Day of the week | 55.01 | 0.0001 | 19.69 |
| leisure/enjoyment difficulties | TST | 12.58 | 0.0004 | 6.69 |
| perceived stress | TST | 14.59 | 0.0001 | 8.7 |
| perceived stress | (DSTP- SSTP) | 6.07 | 0.01 | 0.18 |
| overall satisfaction | TST 2 nights | 10.22 | 0.001 | 4.33 |
| overall ADHD symptom severity | (DSTP- SSTP) | 9.17 | 0.002 | 3.28 |
| BART-Impulsivity | (DSTP- SSTP)^2^ | 20.86 | 0.0001 | 7.65 |
| BART-Impulsivity | TST | 9.47 | 0.002 | 2.86 |

Supplementary Table 2: Outcome of the stepwise modeling procedure and associated statistics

*Final predictors retained for each outcome in the stepwise modeling procedure. For each predictor, we report the likelihood ratio statistic (LR), associated p-value, and change in Bayesian Information Criterion (ΔBIC) relative to the previous model step. Predictors are listed in the order in which they were included in the model (i.e., reflecting their relative contribution to model improvement). Outcomes may appear multiple times when more than one predictor was retained. Family-relationship and peer-relationship difficulty measures are not reported, as no predictors significantly improved model fit for these outcomes.*
